# Resolving RH and GYP hybrid alleles while accessing the entire blood group genome with Nanopore adaptive sampling

**DOI:** 10.1101/2025.06.10.25328638

**Authors:** M. Gueuning, G. A. Thun, S. Koller, S. Sigurdardottir, N. Trost, L. Wagner, B. Mayer, C. Engström, M. P. Mattle-Greminger, S. Meyer

**Affiliations:** Blood Transfusion Service Zurich (SRC), Department of Research and Development, Schlieren, Switzerland; Blood Transfusion Service Zurich (SRC), Department of Molecular Diagnostics and Cytometry, Schlieren, Switzerland; Institute for Transfusion Medicine, Charité– Universitätsmedizin Berlin, Berlin, Germany; Blood Transfusion Service Zurich (SRC), Department of Immunohematology, Schlieren, Switzerland

## Abstract

Adaptive sampling (AS), a computational enrichment method developed for Oxford Nanopore Technologies sequencing platforms, offers a promising advance in molecular blood group diagnostics. By leveraging long-read sequencing, AS has the potential to accurately resolve complex structural variants in the RH and MNS blood group systems, while characterizing the entire blood group genome. In this study, we evaluated the performance of AS using five samples with known or suspected complex variants in the RH and MNS systems, unresolved by standard methods in immunohematology. Samples were sequenced on a PromethION P2 Solo with up to two samples per flowcell, generating 37.0-52.4 Gb of data with mean on-target coverages of 18.9-53.4x, allowing reliable variant detection. Hybrid alleles were characterized using a *de novo* assembly approach, whereas variants in non-recombinant regions were analyzed using both a custom in-house and the EPI2ME reference-based workflow. Considering solely exonic variation, 10-15% of detected alleles contained novel nonsynonymous single nucleotide variants (SNVs) or unreported SNV combinations. All suspected hybrid alleles were successfully assembled and identified as *GYP*401.02, RHD*03N.01*, and *RHD*01EL.44*, representing the first fully characterized haplotypes for these variants publicly available. Overall, AS showed significant potential for advancing blood group genomics by enabling high-resolution, full-gene analysis. Its ability to support high-throughput donor genotyping and precise patient-donor matching may reduce the risk of alloimmunization and delayed hemolytic transfusion reactions, particularly in chronically transfused patients. These findings highlight AS as a powerful tool for both research and clinical applications in transfusion medicine.

## Introduction

Adaptive sampling (AS) represents a considerable innovation in the realm of high-throughput sequencing. This unique feature compatible with Oxford Nanopore Technologies (ONT) sequencing platforms enables enrichment of user-defined genomic regions though a computational approach^1,2^. As DNA strands pass through nanopores, they are immediately mapped against user-defined reference sequences. If a strand does not align with a target region, it is ejected from the nanopore through a reversal of the ionic current, thereby freeing the pore for a new strand. By dynamically selecting genomic regions to be sequenced, AS optimizes sequencing resources and accelerates read acquisition (i.e. sequencing depth) in regions of interests (ROI). In addition to its versatility, the key advantage of this approach is that it eliminates the need for labor-intensive and costly pre-sequencing DNA enrichment procedures.

AS has demonstrated considerable potential in diverse clinical applications, including infectious disease detection^3–5^, rare genetic disorder diagnostics^6,7^, and oncology^8–10^. In blood group genetics, AS holds promise for addressing two major challenges that have proven difficult to resolve using conventional molecular methods.

First, AS allows targeting all the genes relevant for the blood group antigen prediction in a resource-efficient manner, while adhering to ethical guidelines by limiting sequencing to blood group loci. Although clinical transfusion practice typically focuses on a limited subset of blood group systems, comprehensive antigen profiling—encompassing the full erythrocytic antigen profile—is especially advantageous for individuals requiring chronic transfusions. Such patients with conditions like sickle cell disease or thalassemia are at high risk of alloimmunization, i.e. developing antibodies against mismatched blood group antigens from donor blood^11–13^. On the donor side, high-throughput antigen profiling based on array technology has been proposed^14,15^ and is currently implemented in some large blood transfusion centers. However, such approaches have limitations, in particular for the patient side, because they are not comprehensive, as relying on selected genetic variants, and lack flexibility and speed for single sample analysis. By enabling high-resolution, fully phased blood group allele determination for all blood groups, AS offers the potential to extend and facilitate donor-recipient matching, better allocate matched blood units and improve transfusion success rates^16^. This approach is particularly beneficial in populations with diverse genetic backgrounds and, as a consequence, elevated chance of loss of common or formation of uncommon antigens, in which cases identifying antigen-matched donors can be especially challenging^17^.

Second challenge that could be addressed by AS is the accurate characterization of alleles in the RH and MNS blood group systems notorious for harboring complex genetic rearrangements due to the underlying paralogous gene systems (*RHD/RHCE* and glycophorin-encoding genes *GYPA/GYPB/GYPE*, respectively). Although the full genetic resolution of hybrid alleles in the RH and MNS systems is rarely required for routine transfusion decision-making^18^, accurate detection of their presence remains crucial to avoid delayed hemolytic transfusion reactions^19–21^. Most current methods indirectly infer hybrid alleles by relying on selected exonic variants. AS, however, provides two advantages. First, introns are also sequenced allowing breakpoint determination. Second, the long sequenced reads allow assembling adequate reference sequences in order to avoid error-prone variant calling against a reference sequence that considerably deviates from the hybrid allele. Resulting well-characterized hybrid reference sequences will become increasingly valuable for the interpretation of genetic data produced with the advent of high-throughput molecular typing methods in blood group genetics^14,15,22–26^.

In this paper, we examine the potential of AS in blood group genomics, focusing on its ability to resolve hybrid alleles while accessing the entire blood group genome. We outline the advantages but also discuss current limitations hindering its broader adoption and highlight the necessary advancements in blood group-specific bioinformatics tools and reference datasets to further optimize its use.

## Methods

### Pre-Analytics and sample selection

We selected five samples—S01 to S05—with suspected complex genetic variation that could not be fully resolved using standard molecular methods. Four samples were part of the Zurich blood bank donor pool and were all assumed to harbor genetic rearrangements within the RH or MNS blood group systems. One additional sample with undetermined *RHCE* alleles came from a young patient with sickle cell disease requiring chronic transfusions and was provided by the Charité Hospital in Berlin, Germany. Written informed consent for molecular blood group analysis was obtained from all donors, while it was not required for the patient under German national regulations for blood group genotyping.

Standard serological and molecular methods included agglutination testing and genotyping of known causative variation for the most relevant blood group antigens. A detailed description about pre-analytical methods and results are given in supplemental Information Section 1.1 and supplemental Table 1, respectively. In short, S01 was serologically identified as RH:2,-3,-4,5 (CCee), but genotyped as *RHCE*Ce/RHCE*ce* (expecting RH:2,-3,4,5). Neither PCR-SSP kits for RH nor Sanger sequencing of all ten exons suggested causative variation in *RHCE* for the observed phenotype. Similarly, S02 was serologically identified as MNS:1,2 (M+N+) with MNS2 being weakly positive both on automated gel cards system and standard tube testing. Genetic typing and dedicated PCR-SSP kits pointed toward *GYPA*01* homozygosity and suggested the presence of a *GYP*401* (*GYP*Sch*) allele. For S03, the sickle cell patient, anti-e-like and anti-C antibodies were detected, probably from previous transfusions. Sanger sequencing of *RHCE* exons revealed 4 heterozygous single nucleotide variants (SNVs) (c.48G>C; c.105C>T; c.254C>G and c.1025C>T) in addition to the presence of homozygous c.307C (encoding RH:4) and c.676G (encoding RH:5) variants. These SNVs have not yet been described in any combination according to the International Society of Blood Transfusion (ISBT) blood group allele table for *RHCE*^27^. An isolated c.254C>G SNV is defining *RHCE*01.06.01* and the combination of c.48G>C; c.105C>T and c.1025C>T have so far only been described in the presence of c.733C>G and c.744T>C (*RHCE*01.20.04.02*). Haplotype assignment (phasing) of the four heterozygous variants was, however, not possible with the Sanger sequencing data. S04 and S05 both showed positive results for exon 10 in the *RHD*-negative screening, a routinely performed testing for the presence of *RHD* (exons 3, 5 and 10) in RhD-negative first-time donors^28^. Analysis of allelic ratios for exons 3, 4, 5 and 10 in *RHD* vs *RHCE* by MALDI-TOF mass spectrometry (MS) genotyping suggested the presence of hybrid alleles. While S04 showed accordance with a *RHD*03N.01* allele (*RHD-CE(4-7)-D*), S05 pointed towards the presence of *RHD*01EL.44* (*RHD-CE(4-9)-D*) without any RH:1 antigen measurable by flowcytometric analysis or adsorption-elution technique.

### Nanopore sequencing

Genomic DNA was extracted from EDTA whole blood using the Monarch High Molecular Weight DNA extraction kit (New England Biolabs, Ipswich, USA). To control fragment length, the thermal mixer speed was set to 2,000 rpm. Elution was performed in 50 µL Elution Buffer II instead of the standard 100 µL to maximize DNA concentration for library preparation. DNA concentrations were quantified using the Nanodrop 2000 spectrophotometer (Thermo Fisher Scientific, Waltham, USA).

For S01-S03, libraries were individually constructed without barcoding, following ONT’s ligation sequencing gDNA V14 protocol on the PromethION platform (version: GDH_9174_v144_revN_10Nov2022). Because average sequencing coverage with single-sample libraries was ample for reliable variant calling (>35x, Table 1), S04 and S05 were multiplexed to reduce sequencing costs using the ONT Native Barcoding Kit 24 V14 protocol (version: NBE_9163_v225_revQ_15Sep2022).

**Table 1:** Sample-specific sequencing and mapping metrics. Mean coverages have been calculated after basecalling with the fast mode and before excluding reads below a length of 1500 (i.e. ejected reads). Read quality is a measure of accuracy in PHRED score, e.g. 20 and 30 translate to 99% and 99.9% correct base calls.

| Sample | Barcoded | Total Output* (Gb) | N50 (Kb) | Mean Read Quality | Mean sequence length for read classification during AS (± SD) | Mean Coverage (± SD) | |
| --- | --- | --- | --- | --- | --- | --- | --- |
|  |  |  |  |  |  | Inside ROI | Outside ROI |
| S01 | No | 51.7 | 22.0 | 24.7 | 371 (± 82) | 53.4 (± 15.0) | 15.4 (± 8.7) |
| S02 | No | 47.3 | 21.6 | 22.8 | 371 (± 74) | 52.4 (± 15.9) | 15.3 (± 8.0) |
| S03 | No | 37.0 | 18.9 | 23.6 | 370 (± 71) | 38.5 (± 11.8) | 10.9 (± 6.5) |
| S04 | Yes (2x) | 23.6<sup>+</sup> | 25.0 | 22.6 | 361 (± 86) | 27.1 (± 7.6) | 6.5 (± 4.4) |
| S05 | Yes (2x) | 26.0<sup>+</sup> | 32.4 | 22.6 | 361 (± 86) | 18.9 (± 6.6) | 5.0 (± 3.4) |

\*Passing Q8 (minimal quality score)
<sup>+</sup> Total output of 52.4 Gb, including 2.8 Gb that could not be assigned to either S04 or S05.
ROI: region of interest

Libraries were sequenced on a PromethION P2 Solo device using FLO-PRO114M flow cells. Approximately every 24 hours, the flow cells were washed with the Flow Cell Wash Kit (EXP-WSH004) before being reloaded with library. For the ROI in AS, we constructed a FASTA file containing 57 sequences extracted from the latest human reference genome (T2T-CHM13v2.0; accession number: GCF_009914755.1, referred as T2T hereafter). These sequences included 64 target genes (51 encoding Human Erythrocyte Antigens (HEAs), 7 Human Platelet Antigens (HPAs), 4 Human Neutrophil Antigens (HNAs), and 2 transcription factors (TFs)) with the contraction of 13 genes that are located close to each other into six multi-gene loci. Furthermore, a buffer region of 50 kb up– and downstream of the respective genes (loci) was added. Overall, the 57 sequences accounted for 8,680,191 bp representing 0.28% of the total human genome and were on average 152 kb long. A BED file containing the genomic T2T coordinates is provided as supplemental Table 2.

### Computational analyses of Nanopore sequencing data

A flowchart with essential steps in the different workflows is shown in Figure 1.

**Figure 1:**
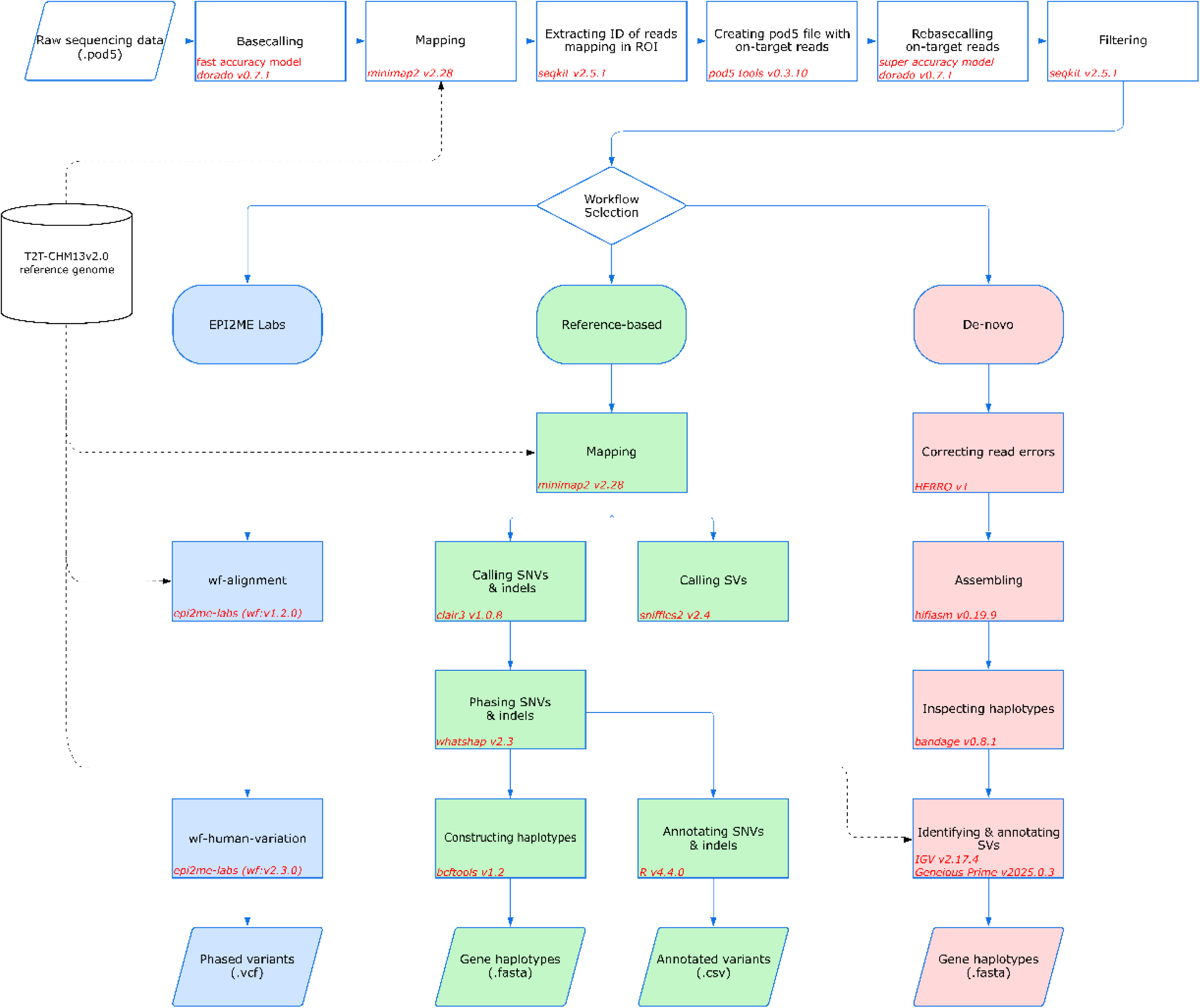
Overview of the bioinformatic workflows applied to ONT sequencing data. A pre-workflow is given at the top of the flowchart before splitting up into the three workflows EPI2ME Labs (blue), inhouse reference-based (green) and *de novo* assembled (red). The tools used at each stage of the analysis are indicated alongside their corresponding step.

### Pre-workflow: Restriction of target reads

First, to minimize computational burden, we initially basecalled and demultiplexed (for S04 and S05) reads using the fast-accuracy model with Dorado (v0.7.1). These reads were subsequently mapped to T2T with Minimap2^29^ (v2.28) using the –m 400 parameter. The resulting SAM file was filtered using SAMtools^30^ (v1.9) to exclude reads with mapping quality scores below 50 or marked as secondary alignments. Converted into a sorted and indexed BAM file, we used Qualimap2^31^ (v2.3) to compute enrichment by comparing coverages in on-vs. off-target regions. The BAM file was also used to calculate mean coverages with mosdepth^32^ (v0.3.9), using a 100 kb window size. Results were plotted using the R packages ggplot2^33^ (v3.5.1) and circlize^34^ (v0.4.16).

From the same BAM file, we extracted read IDs corresponding to sequences overlapping regions specified in the BED file (ROI) using a combination of awk and SAMtools (*samtools view*). The extracted read IDs were then used to filter the original POD5 files containing the electric signal of all reads with ONT’s pod5 tool (v0.3.10). Subsampled POD5 files were rebasecalled and (if applicable) demultiplexed using Dorado’s (v0.7.1) super-accuracy model “dna_r10.4.1_e8.2_400bps_sup@v5.0.0”. BAM files derived from this process were converted to FASTQ format. Finally, reads shorter than 1500 nt or with a quality score of Q<10 were filtered out from the FASTQ files using SAMtools and SeqKit^35^ (v2.5.1), respectively.

### Bioinformatic workflows: Variant calling and haplotype construction

We investigated three distinct bioinformatic workflows for generating haplotypes of blood group as well as HPA and HNA genes: (I) an in-house workflow, (II) ONT’s EPI2ME Labs workflow—both based on reference-based variant calling for all target genes—and (III) a *de novo* workflow for genes with suspected hybrid alleles. A detailed description of each workflow is given in supplemental Information Section 1.2.

Workflow 1. Reads in FASTQ were mapped and filtered for Q≥50 as described in the pre-workflow setting. Variants in the ROI were called with Clair3^36^ (v1.0.8). Resulting calls (SNVs and indels) were filtered with Q10 and phased with WhatsHap^37^ v2.3. For calling structural variants (SVs), defined by a length of at least 50 bp, unmapped reads in FASTQ were first error-corrected by HERRO^38^ (v1), a tool implemented in recent versions of Dorado (v.0.7.1) to generally improve SV calling accuracy. SVs were then called with Sniffles2^39^ (v2.4) and manually phased with SNV data where appropriate. Entire haplotype sequences for all targeted genes with sufficient mean coverage were constructed with BCFtools^30^ (v1.2) by using the variant calls and extracted gene reference data from the T2T genome.

Workflow 2. The EPI2ME Labs workflows “wf-alignment” and “wf-human-variation”, provided by ONT, were downloaded, installed and run with the FASTQ reads with default settings. Using T2T as reference genome, some functionalities like copy number variant (CNV) and short tandem repeat (STR) calling as well as variant annotation were disabled. The aforementioned BED file was used to restrict variant calling to the ROI. Phasing and filtering were carried out as in workflow 1. Final SNV calls from the two workflows were compared with BCFtools.

Workflow 3. To avoid bias from reference sequences, we also assembled FASTQ reads *de novo*. For that, we first corrected reads with HERRO^38^ before applying Hifiasm^40^ v0.19.9. Resulting contigs were visualised with Bandage^41^ v0.8.1 and mapped against T2T. Only contigs of the hybrid alleles of S02 (in MNS), S04 and S05 (in RH) were further inspected by aligning them against publicly available *RHD* and *RHCE* sequences (supplemental Table 3) as well as *GYP(B-A)* hybrid series^42^, which informed breakpoint determination.

SNV and indel calls in workflows 1 and 2 were further restricted with a BED file containing exonic coordinates to output all variants in coding exons or flanking regions (+/-10 nt from exon-intron boundaries). Annotating these variants was carried out with BCFtools and included rs-numbers, consequence on the protein level as well as population-based frequencies. Variants were also manually compared with MALDI-TOF MS genotype data, where available (supplemental Table 4). As ISBT blood group allele tables usually define a dedicated gene reference sequence for each blood group, deviant positions between those and T2T were also inferred (only for HEA and transcription factor encoding genes). Accordingly, reference calls against T2T (genotype 0/0) at such positions were additionally outputted as they differed from the gene reference sequence used by ISBT (genotype 1/1). Because 0/0 calls (reference genotype) often harbor lower calling qualities, their threshold was lowered to Q5 to avoid missing such positions. Finally, all variant calls with respect to the reference sequences used by ISBT were extracted. By using the cDNA annotation with which nucleotide changes are listed in the ISBT allele tables^43^, the two resulting haplotypes per blood group genes were manually compared with the ISBT allele collections. Additionally, we used the tool RBCeq2^44^ to confirm our allele assignments and phenotype predictions. As this tool is based on coordinates of human genome GRCh38, accession number: GCF_000001405.40, we reran EPI2ME Labs (workflow 2) with GRCh38 as reference genome and used the resulting haplotypes as input data for RBCeq2. Coding variants not belonging to reported ISBT alleles were further checked for pathogenicity using CADD^45^ (v1.7) applying PHRED scores for probabilities of altering the phenotype (antigen). Intronic flanking variants were assessed with the splicing module of Alamut(TM) Visual Plus, integrating several splicing prediction algorithms.

Finally, variant calls that were rare in the general population (minor allele frequency, MAF<0.01) were further selected for replication with Sanger sequencing unless they belonged to hybrid alleles implicating a high chance of originating from the paralogous gene. Further details about this procedure are given in supplemental Information Section 1.2.1.

## Results

### Nanopore sequencing output

A detailed description about the sequencing output is provided in supplemental Information Section 2.1. Briefly, PromethION runs yielded 37.0–52.4 Gb of data above the Q8 quality threshold (Table 1), with ∼60% active pores at start. Output was highest for the barcoded run encompassing two samples. Mean on-target coverage per sample ranged from 18.9× to 53.4×, with an average enrichment factor of 3.6 when including all output reads (Table 1). Figure 2A shows coverage across the entire genome for S01 (see supplemental Figure 1 for other samples) and Figure 2B summarizes sample-specific mean coverage per gene with outliers labelled by gene name. *CD99* was excluded from downstream analyses due to consistently minimal coverage across all samples.

**Figure 2:**
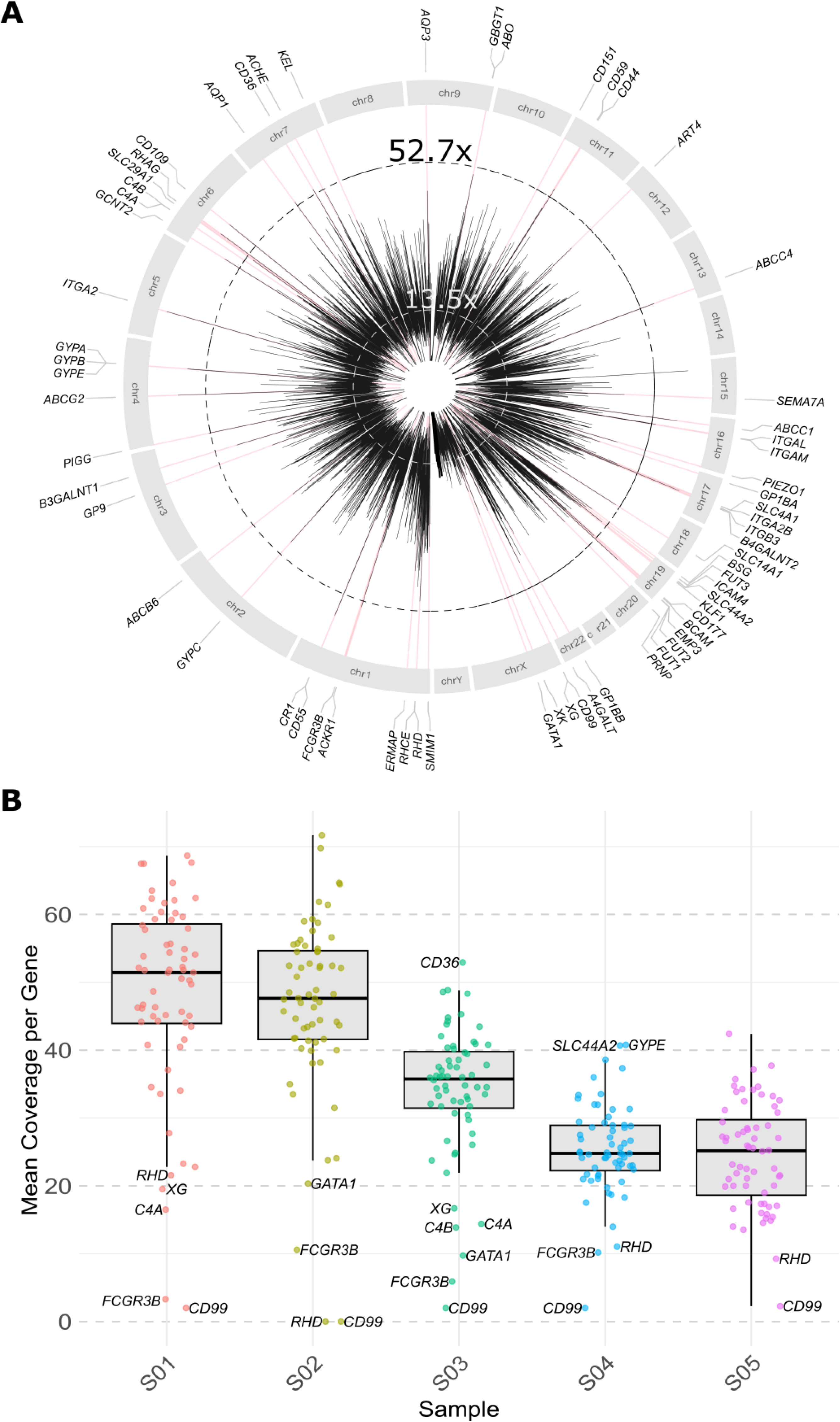
Coverage of on-and off-target regions across samples. **(A)** Circos plot showing read coverage across the genome for S01. Genes of interest are indicated according to chromosomal positions around the outer circle (n=64). Mean coverages across the genome (100 kb windows) is shown by black lines; mean coverages in ROI are further highlighted in red. Circles of dashed lines represent mean read coverage within (black) and outside (white) ROI. Corresponding circos plots for S02 – S05 are shown in supplemental Figure 1. **(B)** Sample-stratified box plots calculated from mean coverages per targeted gene. Genes with coverage identified as outliers or below 10× are labelled by their names.

### Variant calling

The in-house reference-based approach called between 3,778 (S01) and 5,985 (S03) variants within target genes (Table 2), with most variants (>95%) being intronic and ∼85% being SNVs. After filtering to retain only Q10 variants within coding exons (including 10 bp intronic flanking regions), the number of variants dropped to ranges from 130 (S01) to 192 (S05) (Table 2). Due to ambiguous read alignments to *C4A* and *C4B*, as well as to repetitive regions within *CR1* and the *FCGR2*/*FCGR3* gene cluster locus (supplemental Information Section 2.2), we also calculated the number of variants excluding those genes. Variant counts in the rearranged *RHD*/*RHCE* (S04 and S05) and *GYPA*/*GYPB* (S02) hybrid alleles were also omitted (Table 2). Of the remaining variants, nonsynonymous variants were more frequent than synonymous ones in four samples (ratios 1.1 to 1.3), only S03 showed a higher number of synonymous SNVs (ratio 0.7). Between 12 (S04) and 19 (S03) variants were found to lie in flanking intronic regions. However, those predicted to alter spliciSng (supplemental Table 5b) were rare. Frameshift or nonsense mutations were also rare (Table 2) and are discussed in the sample-specific blood group alleles section (see also supplemental Information Section 2.4). Limiting our analysis to blood group genes with defined reference sequences in the ISBT allele tables (n=50), we were able to assign between 58% (S03) and 68% (S04) of all alleles to a corresponding ISBT “star” allele category (Table 3, supplemental Table 6). Among the remaining alleles, between 15% (S04) to 24% (S02) only differed from reported alleles by synonymous SNVs. Prediction of deleteriousness of these variants is given in PHRED scores in supplemental Table 5a. Additionally, we found between 15 and 19 novel alleles harboring at least one nonsynonymous variant and hence with a modified protein sequence (Table 3 and supplemental Table 5a and 6, see also sample-specific blood group alleles). The majority of these nonsynonymous variants were, however, based on unreliable variant calls (supplemental Information Section 2.2) or in alleles of recently discovered blood groups systems (Nr. 37–43, ratified in 2019 or later), which are still lacking comprehensive allele collections. Comparison between our manual allele assignments and those generated by RBCeq2 revealed only few discrepancies (supplemental Information Section 2.3). Final phenotype predictions are given in supplemental Table 7.

**Table 2:** Sample-specific variant calling results. The total number of variants did not include structural variants and excluded any variant calls in *CD99* (Xg blood group system) because of low coverage. Number of variants in coding exons and splice site regions (column 3) were further divided according to their consequences (columns 4 to 8). In brackets, number of variants with high reliability are given. This excludes variants called on *GYPA* and *GYPB* for S02 and those called on *RHD* and *RHCE* for S04 and S05 owing to the presence of hybrid alleles. Due to ambiguous read alignments to *C4A* and *C4B*, as well as to repetitive regions within *CR1* and the *FCGR2*/*FCGR3* gene cluster locus, variants in those four genes were further excluded.

| Sample | Total Nb variants | Variants in coding exons and splice site regions (+-10bp) of ROI (Q10) |  |  |  |  |  |
| --- | --- | --- | --- | --- | --- | --- | --- |
|  |  | Nb variants | Frameshift | Nonsense | Nonsynonymous | Synonymous | Splice Sites |
| S01 | 3778 | 130 **(102)** | 1 | 1 | 59 **(46)** | 50 **(38)** | 19 **(16)** |
| S02 | 4004 | 136 **(108)** | 1 | 1 | 68 **(50)** | 45 **(40)** | 21 **(16)** |
| S03 | 5985 | 172 **(140)** | 1 | 2 | 63 **(49)** | 81 **(69)** | 25 **(19)** |
| S04 | 4439 | 174 **(100)** | 3 **(1)** | 1 | 98 **(45)** | 56 **(41)** | 16 **(12)** |
| S05 | 4309 | 192 **(120)** | 2 **(1)** | 2 | 110 **(57)** | 61 **(44)** | 17 **(16)** |

**Table 3:** Sample-specific summary of assigned blood group alleles. Included were 45 blood group systems recognized by ISBT (as of May 2024), but the last ratified system, CD36, had to be excluded since no official reference sequence was yet defined. Due to low coverages across samples, *CD99* allele determination (Xg blood group) was also excluded. Reference-matching alleles are those, with respect to coding variation, identical to the ISBT-defined reference allele for each blood group gene (e.g. *ABO*A1.01* for *ABO*). Results of the pathogenicity analysis of variants in novel alleles are provided in supplemental Table 5. Due to ambiguous read alignments to *C4A* and *C4B*, as well as to repetitive regions within *CR1* and the *FCGR2*/*FCGR3* gene cluster locus, allele assignment for these genes is less reliable and number of alleles excluding those genes are given in brackets.

| Sample | Assigned Alleles* | Reference-Matching Alleles | Other Known Star Alleles | Novel alleles with only Synonymous variants | Novel alleles with Syn+Nonsynonymous Variants |
| --- | --- | --- | --- | --- | --- |
| S01 | 97 | 53 | 9 | 20 **(19)** | 15 **(10)** |
| S02 | 97 | 49 **(48)** | 9 | 23 | 16 **(11)** |
| S03 | 97 | 47 | 9 | 22 | 19 **(13)** |
| S04 | 100 | 55 | 13 | 15 | 17 **(11)** |
| S05 | 97 | 50 | 10 | 21 **(20)** | 16 **(11)** |

\*Include only ISBT-ratified blood groups with official reference genes as of May 2024 (i.e. excluding *CD36*). *CD99* was excluded due to consistently minimal coverage across all samples. Differences in number of assigned alleles are due to blood group genes located on the X chromosome (i.e. *XK*, *XG* and *GATA1*).

### Verification of rare variants

With the in-house workflow, we identified 78 variants located in exonic and flanking intronic regions which were rare (or not annotated) in large whole genome sequencing (WGS) studies. Among these, 13 were shared between two samples (supplemental Table 8). Five *GYPA* variants in S02 were explained by the *GYP*401* hybrid allele and were excluded. Of the remaining variants, 47—including all those detected in two individuals—were excluded due to their location in *C4A*/*C4B*, *CR1* or *FCGR3B,* see supplemental Information Section 2.2 and 2.4. The remaining 26 variants (25 SNVs and one indel), distributed over 18 genes, were all successfully confirmed with Sanger sequencing.

### Structural variant calling

The number of SVs called in the in-house workflow ranged from 18 to 33 per sample and these were exclusively insertions and deletions (supplemental Table 9). The median lengths of insertions per sample (110 to 330 bp) was comparable to the lengths of deletions (109 to 210 bp). Only two deletions were shared across all samples. One 192 bp deletion rs71151470 in intron 14 of the integrin-encoding *ITGAM* gene (NC_060940.1:g.31703459-31703651) and the 109 bp deletion rs1557633501 associated with RH4 expression of *RHCE* (NC_060925.1:g.25241875-25241984). SV calling tool Sniffles2 failed to identify the large deletions associated with *RHD*01N.01* (i.e. the loss of the entire *RHD* gene) present in all the samples. Additional filtering to retain only SVs overlapping with coding exons excluded all variants in S02 to S05. For S01, a single deletion of 8,636 bp spanning *RHCE* introns 8 and 9 remained (NC_060925.1:g.25199922–25208558), see sample-specific blood group alleles. We confirmed this deletion using a bridge-PCR approach (supplemental Information Section 1.2.1 and supplemental Figure 2).

### Comparison between in-house and EPI2ME Labs workflows

Variant calling (SNVs and indels) using the two reference-based workflows produced highly similar results. Restricting the comparison to Q10 variants in coding regions revealed identical calls for S01 and S05. For the other three samples, differences were minimal. Specifically, the EPI2ME Labs workflow identified three additional variants in S02, two additional variants while missing one in S03; and called two fewer variants in S04. Further investigation showed that all these discrepancies concerned genes, in which variant calling was regarded as unreliable (*GYPA* in S02; *C4A*, *C4B*, *FCGR3B*, see supplemental Information Section 2.2).

### De-novo assembly of hybrid alleles

As an alternative to haplotyping based on reference sequences (workflow 1 and 2), we *de novo* assembled sequencing reads for the 57 ROI (supplemental Table 2), producing per sample between 103 and 156 primary contigs (i.e. haplotype consensus sequences). N50 values ranged from 136 (S03) to 222 kb (S02) (supplemental Table 10). Knowing that our target loci were on average 152 kb, these results indicate a relatively high contiguity of the assemblies. A detailed analysis of all inferred haplotigs (contigs coming from the same haplotype) was beyond the scope of this work, but we focused on those representing the suspected hybrid alleles. Importantly, the presence of long contigs for the locus encoding MNS antigens (S02) and RH antigens (S04, S05) allowed resolving all complex genetic rearrangements investigated in this study (see sample-specific blood group alleles).

### Sample-specific blood group alleles

Here, we focus on the most relevant blood group alleles identified in each sample— specifically those that could not be fully resolved using alternative technologies. Summaries of additional findings for each sample are provided in supplemental Results 2.5. For hybrid alleles, we used the *de novo* assembly workflow, given the high likelihood of ambiguous read mapping and the frequent failure of SV callers to detect these recombination events.

S01 was the only sample harboring a SV spanning coding exons. We identified a novel 8,636 bp deletion spanning from *RHCE* intron 8 to 9. This represents the largest deletion reported in the *RHCE* gene to date. The deletion was on the haplotype carrying the *RHCE*01* allele and explains the original phenotype-genotype discrepancy, i.e. the absent RH4 expression. Moreover, this deletion was not only called in the SV calling step of the reference-based workflows, but also identified through the *de novo* assembly workflow where it was present within a 254.7 kb contig, spanning the entire *RH* locus composed of *RHD*01N.01* (i.e. absence of *RHD*) and *RHCE*01* (Figure 3).

**Figure 3:**
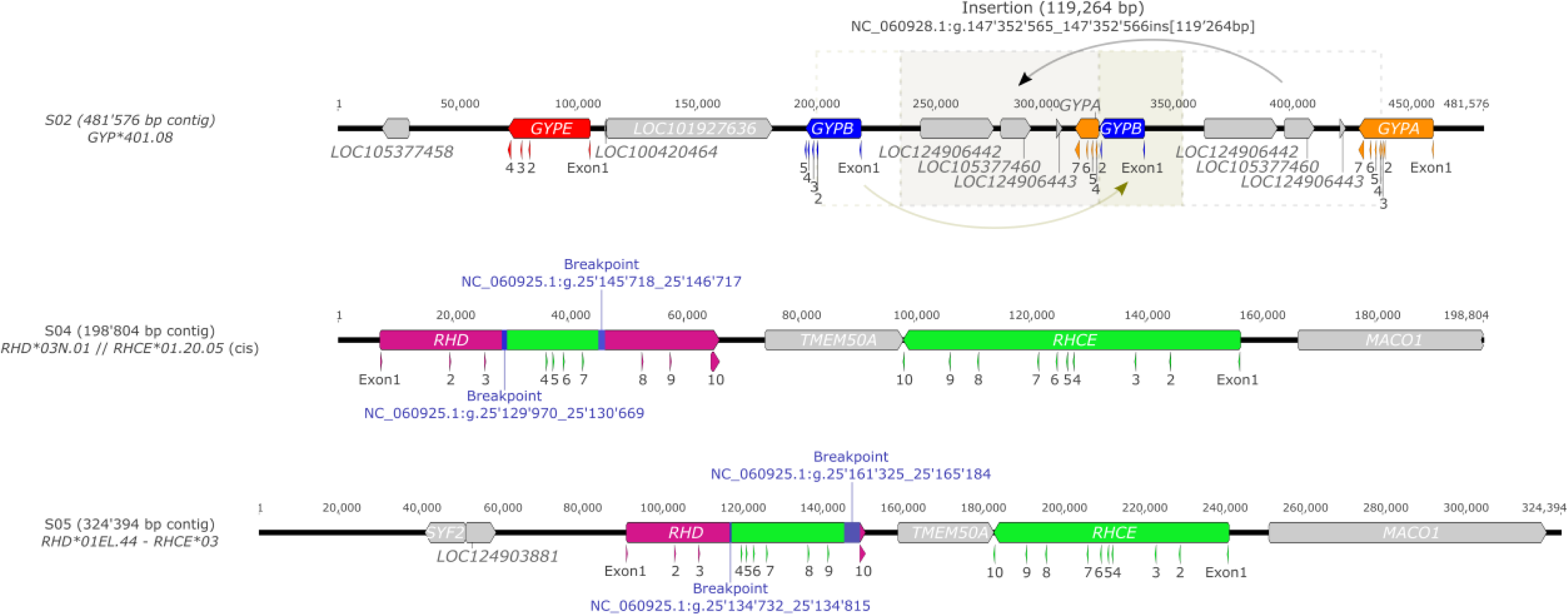
Overview of contigs containing hybrid alleles. Contigs were assembled using Hifiasm (v0.19.9) and annotated with a BLAST-like tool integrated into Geneious Prime (v2025.0.3), using official ISBT blood group reference annotations. Contig lengths were preserved as originally assembled.

For S02, the *de novo* workflow assembled a 481.6 kb contig, encompassing *GYPA*, *GYPB*, and *GYPE*, as well as including a 119,264 bp insertion at NC_060928.1:g.147352565, Figure 3. This insertion contained a hybrid gene formed between *GYPB* (exons 1, 2 and pseudoexon 3) and *GYPA* (exons 4–7), corresponding to the *GYP*401* allele types, which explained the weak MNS2 reaction without the presence of a *GYPA*02* allele. Mapping to *GYPA* and *GYPB* reference sequences (NG_007470.3; NG_007483.2) allowed narrowing down the recombination site between *GYPB* and *GYPA* in the insertion to ∼150 bp. Comparison with all ten *GYP*401* sequences listed in the ISBT allele table^42^ showed identical breakpoint region to *GYP*401.02*, also called *GYP*Sch(type B)*, with only two divergent nucleotides in the ∼940 bp intronic region between pseudoexon 3 and exon 4^46,47^. Both divergent SNVs originated from *GYPA*. One, at position 696, was an unknown SNV shared only between the *GYP*401.01* and *02* reference sequences and could potentially be a sequencing error. The second divergent SNV, at position 776, was a common *GYPA* variant (rs115171794). While the breakpoint region of our assembled allele was also in agreement with *GYP*401*.*10*, we could not further investigate similarity due to the absence of a continuous sequence for that allele. This allele, however, deviates from *GYP*401.02*–and our assembled sequence–at position 774^47^. Our assembly also revealed *GYPA*01* and *GYPB*04.06* alleles as part of the *GYP*401.02* haplotype. These two alleles were also detected *in trans*.

In S03, detected exonic *RHCE* variants by Sanger sequencing could previously not be assigned to separate haplotypes (supplemental Table 1). Our variant calling workflow phased variants c.48G>C, c.105C>T and c.1025C>T to the same haplotype, forming a new ISBT allele on the basis of *RHCE*01.02.01* with additional c.105C>T (p.Asp35=), which is not expected to change phenotype (i.e. weak/partial RH4 and RH5). As for the other haplotype, c.254G>C was the only variant called and defines the *RHCE*01.06.01* allele, which modifies antigen RH5. With respect to MNS, we detected the null allele *GYPB*03N.04* in combination with synonymous c.96T>C and c.102A>G. This allele included the exon 5 skipping c.270+5G>T variant (rs139511876^48^), which was reassuringly predicted to essentially weaken the splice site between exon and intron 5 (supplemental Table 5b). The MNS:-3,w5 phenotype predicted for this allele was serologically masked by *GYPB*03* on the other haplotype, but a minimal MNS6 expression (He+) has been described^49^.

For S04, as suspected by results from MALDI-TOF MS pre-analyses, we identified a *RHD-CE-D* hybrid allele located on a 198.8 kb-long contig. The allele comprised exons 1–3 and 8-10 from *RHD*, separated by *RHCE* exons 4–7. Additionally, this contig exhibited c.186T in exon 2 as well as c.410T and c.455C in exon 3–all characteristic of the *RHD*03N.01* allele–which explained the weak RH2 positivity in the absence of any *RHCE*02* allele (RH10 and RH20 were not determined). This hybrid allele was *in cis* with *RHCE*01.20.05*, defined by c.733G and c.1006T positions, which were also found in exons 5 and 7 within the hybrid. Mapping 25 publicly available full or partial *RHD* and *RHCE* sequences (supplemental Table 3) to our contig narrowed down potential breakpoints to stretches of a few hundred base pairs in introns 3 (NC_060925.1:g.25129970-25130669) and 7 (NC_060925.1:g.25145718-25146717). The identified breakpoints were consistent with those previously described^50^.

For S05, we detected a *RHD-CE-D* hybrid allele located on a 324.4 kb contig. The part with *RHD* origin comprised exons 1–3, followed by exons 4–9 from *RHCE*, and exon 10 from *RHD,* characteristic of *RHD*01EL.44*. The hybrid allele was *in cis* with *a RHCE*03* allele. Analysis of breakpoints using the same strategy as for S04 identified a homologous region in intron 3, where recombination likely occurred. We could narrow down this 5’ breakpoint region to fewer than 100 bp (NC_060925.1:g.25134732-25134815), which closely resembled the one reported by Srivastava and colleagues^51^ (accession number: OP804519). However, in intron 9, the breakpoint region was larger (∼4 kb) due to identity of the two genes (NC_060925.1:g.25161325-25165184). Although this allele is classified as *RHD*01EL*, causing a Del (D-elute) phenotype, we could not detect any D antigen, neither with adsorption-elution (performed on two independent samples) nor with flow cytometry techniques.

## Discussion

In this study we evaluated the effectiveness of AS in resolving hybrid alleles in the RH and MNS blood group systems while in parallel assessing its ability to provide complete and phased genetic information for all known blood group genes.

### Resolving hybrid alleles

Using a *de novo* approach primarily based on the assembly tool Hifiasm with prior read error correction with HERRO, we were able to construct haplotype contigs ranging from 199 to 482 kb, spanning the entire *RH* or *GYP* locus with their respective hybrid alleles. For the two investigated *RH* hybrid alleles (S04: *RHD*03N.01* and S05: *RHD*01EL.44*), breakpoint regions were comparable to those previously reported^50,51^. Due to the high sequence homology between *RHD* and *RHCE*, breakpoints can often not be exactly determined. Since particularly the latter half of intron 9 is identical between *RHD* and *RHCE* reference sequences, we were only able to approximate the breakpoint initiation site for *RHD*01EL.44* to a ∼4 kb region. Notably, both adsorption-elution and flow cytometric testing on S05 yielded negative results, which differed from previous in-house experience with this hybrid type. While small differences in the 5’ breakpoint could be responsible for this finding, DEL phenotypes may also misclassify as D-negative when expressing a partial phenotype^52^. Regarding the *GYP* hybrid allele of sample S02, we identified a ∼150 bp region in the insertion where recombination between *GYPB* and *GYPA* likely occurred. Nevertheless, exact breakpoint determination also proved in this case infeasible due to high sequence similarity. To the best of our knowledge, this is the first *GYP*401* allele spanning the entire locus that has ever been reported, highlighting the critical need for publicly accessible full haplotype sequences to facilitate accurate characterization and validation of similar sequences.

Hybrid alleles have long occupied the blood group genetics community. Approaches based on short read sequencing of exons had some success by inferring hybrids based on RH (and GYP) exon coverage ratios^53–56^. Short read data from WGS studies or from long-range PCR enriched RH amplicons allowed even full-gene haplotyping of hybrid alleles, but coverage ratios remained delicate to interpret and phasing uncertainties could not be fully eliminated^57–60^. With the advent of accurate long read sequencing, unambiguous read mapping in repetitive regions became feasible. Nevertheless, current SV callers developed for long read data still struggle with translocations in paralogous gene systems. This challenge is evident in our study, where the SV calling tool not only failed at calling the translocations (hybrid alleles), but also missed the deletions of the *RHD* gene in all the samples despite the availability of relatively long reads (N50 > 18.9 kb).

As a consequence of the limited success of these reference-based approaches, assembly-based methods have emerged. We used here the specific combination of Hifiasm and HERRO, which was critical for achieving high-quality assemblies, as alternative assemblers or omission of the read correction tool resulted in shorter, fragmented, and incomplete contigs. Similar, but less generalizable methods include capture-based enrichment of RH sequences resulting in 2-3 kb long consensus fragments sequenced by PacBio SMRT sequencing^61^. Assembly success was, however, dependent on a high heterogenicity of the samples owing to the modest read length. A similar tool (Chinook), also based on capturing target genes followed by a locus-specific assembly procedure was recently introduced by ONT, but is currently limited to resolve complex SVs in the *CYP2D6/D7/D8* gene-pseudogene system. Of note, we observed three more loci, for which reference-based variant calling failed. *C4A/B*, *CR1* and *FCGR3B* would all require assembly-based strategies to reliably disentangle the allelic composition (which was beyond the scope of this study). In future, replacing single reference genomes by pangenomes alleviate the dependency on the representativeness of the chosen reference genome and will help reducing erroneous variant calls, particularly at complex genomic loci^62^ (See supplemental Information Section 3.1).

### Resolving the entire blood group genome

Extensive blood group characterization has recently become a primary goal for many large blood transfusion centers. It has the potential to reduce the risk of alloimmunization and delayed hemolytic transfusion reactions in chronically transfused patients, but it also enables blood banks to optimize inventory by efficiently managing rare or antigen-matched blood units. Consequently, there has been growing interest in developing high-throughput genotyping methods for both patients and donors, either by MALDI-TOF MS technology^63,64^, by BeadChip arrays^65^ or even high density GeneChip arrays^15,66^. While these methods are cost-efficient and accurate, inferring blood group alleles from genotype data is incomplete and relies on statistical phasing, i.e. on estimating the maternal and paternal haplotype with the help of large population-based genotype data. This is particularly error-prone if variants are rare, in regions with low genetic complexity or low linkage disequilibrium, or if structural variants are present. The challenge is further amplified in genetically diverse or underrepresented populations, where large-scale reference data is missing.

Specific sequencing of blood group genes is a complementary, more comprehensive approach, although currently not intended to replace high-throughput genotyping methods due to costs. So far, it has typically been done in the form of targeted exome sequencing^54^ by which information in coding regions is complete, but phasing uncertainties and incomplete haplotypes remain. AS, as it is based on long-read sequencing, solves these issues, and we produced full-gene and fully phased haplotypes for ∼50 loci. While the relatively high costs, currently ∼$600/sample when barcoding 2 samples per flowcell, makes it impractical for routine use, AS is particularly beneficial for cases involving unexplained discrepancies in the RH or MNS system, for frequently transfused patients or may also be helpful for urgent clinical scenarios where rapid turnaround is critical. For example, in this study, AS was applied to a young patient with sickle cell disease (S03), which could resolve allelic composition for all blood groups including phased–*RHCE* alleles with an uncharacterized constellation of SNVs explaining RH:5(weak/partial) and the presence of anti e-like and anti-C antibodies. Given the patient’s young age, having a complete blood group genome will facilitate ideal donor matching, thereby reducing the risk of further alloimmunization at young age. However, to maximize the benefits of such comprehensive blood group resolution, it is essential to have equivalent genotyping or sequencing methods running in parallel to provide extended genotype data for a large and diverse panel of donors. This integration of AS-based sequencing with scalable genotyping approaches will be key to improving transfusion safety and advancing personalized blood matching strategies in the future.

Full-gene haplotypes also provide comprehensive genetic information in often overlooked regions such as intronic and promoter regions, along with their associated regulatory elements^67–69^. Although the regulation of many blood group genes remains poorly understood, a growing body of literature indicates that variants in these regions can dictate differential gene expression^70–72^. Currently, only ∼130 intronic variants—primarily located in splice sites—are documented in the ISBT allele tables^43^. However, this number is expected to rise with the growing number of sequencing studies. In our analysis of only five samples, we identified two previously unreported variants predicted to form alternative splice sites (in *ERMAP*, SC blood group, and *B4GALNT2*, SID blood group). Furthermore, we detected at least 56 novel alleles based on nonsynonymous variation, not considering the blood group genes with unreliable variant calling, i.e. one in eight alleles harbored novel coding SNVs or unreported SNV combinations (Table 3). Notably, almost half of these novel alleles were found in genes of recently ratified blood groups with relatively low clinical significance (supplemental Table 6).

This continuous discovery of novel alleles presents new challenges for the curation and classification of blood group alleles. Moreover, the ability to resolve full-length haplotypes will probably become a new standard, requiring updates to existing nomenclature guidelines, including a revision of what constitutes a unique blood group allele. A classification system based solely on exonic and splice-site variants is no longer sufficient to reflect the rapid accumulation of haplotype-resolved sequences covering entire genes^58,59,68,69,73^.

### Further advantages compared to other long-read enrichment methods

As exemplified in the last two paragraphs, AS excels in constructing full haplotypes for complex hybrid alleles while simultaneously characterizing the entire blood group genome, AS has some other none neglectable advantages. Other long-read enrichment methods have been applied, including long-range PCR^68^, Cas9-guided adapter ligation^74^, hybridisation-based long-read capturing^75^. While the former two are not suitable to target several genes in parallel, the capture-based method has already been successfully applied to the blood group genome^75^. Read length limitations (mean: 6.3 kb ± 0.3 kb), however, may be a caveat to successfully apply assembly-based methods to resolve hybrid alleles. Major advantages of AS are the fast and easy step to setup-up target regions by simply adding or deleting target loci in the BED or FASTA file. Furthermore, AS does not require any molecular enrichment meaning that the protocol from extracted genomic DNA to first sequences can be performed in approximately two hours. Finally, Nanopore sequencing enables real-time data generation, permitting analysis even before the sequencing run is complete—a valuable feature in urgent clinical settings requiring rapid and precise blood group determination. The only other option with the same capabilities in terms of speed, comprehensiveness and read length is Nanopore WGS, a strategy which has already been used to resolve a complex hybrid allele^76^. Beside being at least 4x more expensive, such a procedure for clarifying blood group genes is not practical as ethically controversial in Switzerland or Germany and would also require extended informed consent by the donor or patient.

### Limitations and challenges

The primary limitation to broader adoption of AS remains its relatively high per-sample cost. However, ongoing improvements in sequencing chemistry and increased data yield are making higher levels of multiplexing feasible, which is expected to significantly reduce costs in the near future. A second barrier to widespread use of AS has been the technical expertise required, both in terms of lab work and bioinformatics. However, recent improvements in ONT’s chemistry, protocols, and user-friendly tools like EPI2ME Labs have significantly lowered this hurdle. In our study, EPI2ME Labs delivered reliable variant calling results with only marginal differences in comparisons with our custom workflow. One primary remaining challenge lies in translating variant calls into blood group alleles. Although a few commercial tools exist for converting variant call format (VCF) files into ISBT “star” alleles (e.g. bloodTyper^77^ from GenomeSys or NGStype software from Innotrain), they are specifically designed for short-read sequencing data and do not currently support the integration of fully phased variants across entire genes. More recently, a tool based on machine learning has been introduced for blood group prediction when large-scale genotype data is available^78^. Finally, the software RBCeq2 announced a first release when this manuscript was drafted, and a first check with our data was successful.

In summary, Nanopore sequencing of the blood group genome, targeted with adaptive sampling, proved highly suitable to elucidate allelic composition in all blood group systems. Complex hybrid alleles in the RH or MNS systems were successfully resolved by the choice of an assembly-based workflow.

## Acknowledgments

This work was financially supported by the Blood Transfusion Service Zurich SRC (Switzerland).

## Authorship Contributions

Contribution: M.G., GA.T., MP.M.-G. and S.M. initiated the study and contributed ideas; M.G., GA.T., MP.M.-G. and S.M. designed the study and experiments; S.S., N.T., L.W., B.M and C.E. contributed samples and provided pre-analysis data; M.G., GA.T. and S.K. performed experiments and analysed data; M.G. and GA.T. wrote the manuscript; all authors commented on the manuscript and approved the final version.

## Data availability

All haplotype sequences can be obtained from the NCBI GenBank sequence database (accession numbers: XXX-XXX). A detailed list of sequence accession numbers is provided in supplemental Table 11 (in submission).

