## Supplemental Information for "Resolving RH and GYP hybrid alleles while accessing the entire blood group genome with Nanopore adaptive sampling"

**Affiliations:**

Morgan Gueuning, Department of Research and Development, Blood Transfusion Service Zurich, Swiss Red Cross, Rütistrasse 19, 8952 Schlieren, Switzerland

### Section 1: Material and Methods

#### 1.1 Pre-Analytics and sample selection

All donor samples were serologically tested for ABO, RH (RH1-RH5, RH8) and KEL (KEL1) antigens using an automated NEO Iris platform (Immucore, Rödermark, Germany). Being regular donors, S01 and S02 were additionally run on an Erytra automated gel cards system (Grifols, Barcelona, Spain) typing for 16 antigens (k, Kpa, Kpb, Fya, Fyb, Jka, Jkb, Lea, Leb, P1, M, N, S, s , Lua, Lub) in 7 systems (KEL, FY, JK, LE, P1PK, MNS, LU). The patient's blood samples were routinely phenotyped using the automated NEO Iris system, and the extended phenotype (FY, JK, MNS) was determined using gel cards on the Erytra platform. The patient was further genotyped for *RHD* and *RHCE* variants using real-time PCR with the ERY Q RH kit (BAG, Lich, Germany). All samples were genotyped on high-throughput MALDI-TOF mass spectrometry (MS) assessing 29 single nucleotide variants (SNVs) spread across 15 blood group systems accounting for 45 human erythrocyte antigens (HEA) and two platelet systems with 4 human platelet antigens (HPA)[1]. Being part of a special sample pool, S01 was further genotyped on 42 additional SNVs amounting the number of blood group systems to 17. Due to observed incongruities, the donor samples were further analysed with commercial (Inno-train, Kronberg, Germany) and/or in-house PCR-SSP kits (sequence-specific-priming PCR). For S05, we further investigated the presence of RHCE antigens on the erythrocytes using flowcytometric analyses. Additionally, the presence of RHD antigens were further analysed by the adsorption-elution method[2]. Sanger sequencing of the *RHCE* exons was carried out for the patient (S03) as well as for donors S01 and S04.

#### 1.2 Bioinformatic workflows

##### 1.2.1 Worflow 1: variant calling

For the inhouse reference-based workflow (I), Figure 1, we started by mapping the filtered FASTQ reads called with the super-accuracy model to the human reference genome (T2T-CHM13v2.0; accession number: GCF_009914755.1, in short: T2T) using Minimap2[3] (v2.28) with the -m 400 parameter. SAMtools[4] (v1.9) was then used to filter out secondary alignments and reads with mapping quality below 50, before being converted to a sorted and indexed BAM file. Variants were subsequently called using Clair3[5] (v1.0.8) in genomic regions containing our genes of interest. The variant calling model was adjusted according to the basecalling model (r1041_e82_400bps_sup_v500). Furthermore, variant calling threshold was set to Q10 without specifying a minimal alternative allele frequency threshold for single nucleotide variants (SNVs) and indels. We also applied the "print reference calls" option to obtain a genomic VCF file containing all positions rather than only the ones different from the reference genome. Variants including SNVs and indels were then phased with WhatsHap[6] (v2.3). The resulting VCF file was additionally filtered using VCFtools[7] (v0.1.16) to only retain variants present in coding sequences (CDS) using a BED file containing genomic coordinates of all coding exons. To avoid filtering out splice site variants, we added 10 bp upstream and downstream of each coding exon.

Structural variants (SVs) were called with Sniffles2[8] (v2.4). To improve calling accuracy we utilized error-corrected reads generated by HERRO[9] and mapped against T2T using Minimap2 with the same parameters as described above. The resulting BAM files were processed with Sniffles2, with all parameters set to their default values. The output VCF files were subsequently filtered using the BCFtools'[4] (v1.2) *intersect* function to retain only SVs present within the target genes, followed by an additional filtering step to include only those overlapping coding exons. To assess potential overlap of SVs among the five samples, the BCFtools' *isec* function was used. Phasing with short variant data was done manually.

*Annotating variant calls and comparison with relevant allele collections*

For downstream analyses, we added rs-number annotations to the phased VCF files using a dbSNP lift over file linking variant coordinates from the human genome GRCh38, accession number GCF_000001405.40, to the T2T assembly using the annotate function implemented in BCFtools. Variants were further annotated and filtered using a custom R script to compute amino acid changes for nonsynonymous variants. For that, genomic annotations from the T2T reference genome were imported as a GFF file and processed to create a transcript database (TxDb) for extracting transcript IDs and names. Transcripts of interest were selected from International Society of Blood Transfusion (ISBT) allele tables[10] for human erythrocyte antigens (HEA). A BED file specifying these transcripts was used to refine the analysis to relevant regions. Using the "predict coding" function from the VariantAnnotation package[11] (v1.50.0), coding variants were computed by integrating the VCF data with transcript and genomic sequence information. After filtering steps to keep only variants tagged as "PASS" (i.e. Q>=10), additional annotations were added, including T2T-specific differences compared to the gene reference sequences used by ISBT, usually corresponding to Locus Reference Genomic (LRG) sequences, as well as minor allele frequencies (MAF) of several population-based whole genome sequencing (WGS) studies obtained via the R package RSNPS[12]. The resulting data were organized into comprehensive tables used for manually assigning star alleles for all blood group genes based on ISBT allele tables. For HPA and HNA systems, we did not assign star alleles because curated allele collections were only partially available[13,14].

*Replication of rare variants*

Variants considered rare in the two largest population-based WGS studies (MAF<0.01 in GnomAD[15] or TopMed[16]) were verified using Sanger sequencing. In case of missing frequencies in both studies, frequencies in further studies (1000G[17], GnomAD-exome[18] and dbGaP) were consulted and SNVs excluded if MAF was above 0.01 in at least one of the studies. Remaining rare variants present in at least three of the five individuals were manually checked and could all be excluded due to either flipped allele assignment or faulty frequencies in the general population due to paralogous gene systems. We further excluded variants located in the expected hybrid allele due to high chance of representing the paralogous gene, i.e. called owing to mismappings. Primers for Sanger sequencing were designed using Primer3[19] (v2.3.7), targeting regions within 500 bp upstream and downstream of each variant. The SV detected in S01 was further investigated by designing gap-PCR assay. One primer pair was designed to span the flanking regions of the deletion, with the forward primer positioned upstream of the 5′ breakpoint and the reverse primer downstream of the 3′ breakpoint. To assess the zygosity of the deletion, an additional primer was designed within the deleted region and paired with the upstream forward primer. In total, three primer combinations were tested, each validated against three wild-type control samples.

*Construction of full-gene haplotypes*

For all samples, we constructed haplotype sequences for up to 63 target genes with mean coverage >10x. For this, the phased VCF files were first hard filtered for variants not passing the quality threshold (i.e. Q<10). In parallel, SAMtools' *faidx* was used to subsample the T2T reference genome to our genes of interest. This subset reference genome was then used in combination with the filtered VCF file to output both haplotypes per gene using BCFtools' *consensus* tool. Haplotypes were then annotated with CDS annotations from T2T using a blast-like tool integrated in Geneious Prime (v2025.0.3). Haplotypes for hybrid alleles were exempt from this procedure and were instead constructed using a *de novo* approach (see below). For the three genes on the X-chromosome, only one haplotype was generated for the male samples S01, S02, S03 and S05.

##### 1.2.2 Workflow 2: variant calling

The EPI2ME Labs workflow (II), Figure 1, involved aligning the filtered FASTQ reads to T2T using the alignment workflow (v1.2.0) with default parameters. The resulting BAM file was then processed through the human-variation workflow (v2.3.0) to call SNVs, indels and SVs. As the T2T reference genome was employed, the options for calling copy number variants (CNVs), short tandem repeats (STRs) and annotations were disabled. The "-include_all_ctgs" option was set to true, the minimum coverage threshold was set to 7 and phasing was enabled. The "-override_basecaller_cfg" parameter pointed to the correct basecalling model used, and a BED file delineating target genes was provided to restrict variant calling to these specific regions. As described in the first workflow, VCFs were finally filtered to keep only variants present in coding regions with a quality score equal or above 10. Variants were also annotated with rs-numbers like in the inhouse reference-based workflow. Variants within coding regions were compared between workflows (I) and (II) using the BCFtools' *isec* function.

##### 1.2.3 Workflow 3: De-novo assembly of gene haplotypes

A *de novo* workflow (III), Figure 1, was specifically used for resolving hybrid alleles. First, the filtered FASTQ reads were corrected using HERRO implemented in Dorado (v0.7.1). Resulting corrected FASTA reads were assembled with Hifiasm[20] (v0.19.9) with the "purging duplicates" option disabled (-l 0). Contigs were then visualized in Bandage[21] (v0.8.1) before being exported in FASTA format. Summary statistics were generated by SeqFu[22] (v1.20.3). All contigs were mapped against T2T using Minimap2. Only contigs corresponding to hybrid alleles were further inspected and annotated. To determine the exact *GYP*401* (*GYP*Sch* type) allele, we downloaded all sequences listed in the ISBT allele table[23] that are associated to a *GYP*Sch* type and aligned them with Minimap2 to the corresponding contig. Similarly, to identify potential breakpoint regions for the two *RHD* hybrid alleles (S04 and S05), we downloaded and aligned 25 publicly available *RHD* and *RHCE* sequences using Minimap2 (sequence accession numbers are provided in supplemental Table 3). By manually inspecting the alignments, we identified and differentiated regions homologous to *RHD* and *RHCE*, respectively.

### Section 2: Results

#### 2.1 Nanopore sequencing output

PromethION runs generated between 37.0 and 52.4 Gb of data passing the default Q8 read quality threshold implemented in MinKNOW, the operating software for Nanopore sequencing devices (Table 1). Flow cells exhibited between 57.9 and 63.5% active pores at start. The barcoded sequencing run yielded highest total output despite 5.4% unclassified reads. The N50 values varied from 18.9 kb to 32.4 kb per sample, with the highest values observed in the barcoded run. Analysis of the adaptive sampling metrics revealed that read classification as on-target or off-target was very consistent across the four runs, with mean decision times per run of approximately 370 (±80) sequenced nucleotides (Table 1). Mean coverage for on-target regions per sample ranged between 18.9 (±6.6) and 53.4 (±15.0), while coverage for off-target regions ranged from 5.0 (±3.4) to 15.4 (±8.7), corresponding to an average enrichment factor of 3.6 (Table 1). Figure 2A and supplemental Figure 1 display mean read depth in 100 kb windows for both on-and off-target regions. Values per gene of interest are given in Figure 2B where genes with read depths classified as outliers (values falling outside first quartile minus 1.5* interquartile range (IQR) or third quartile plus 1.5*IQR) or with coverage below 10x were labeled by their gene names. Ten genes (*C4A, C4B, CD36, CD99, FCGR3B, GATA1, GYPE, RHD, SLC44A2, XG*) were outliers in at least one sample. Three were slightly overrepresented (*CD36*, *SLC44A2* and *GYPE*), whereas the other seven genes were underrepresented. *C4A* and *C4B* are highly homologous (see Section 2.2), which could lead to uneven read assignment and thus coverage reduction in one of the genes. *FCGR3B* had low coverage in S01–S04, likely attributed to high homology in this region (Section 2.2). In fact, the *FCGR2/3* locus is known to frequently harbor CNVs, and the T2T reference genome includes an additional segmental duplication (CN=2) compared to the GRCh38 genome (CN=1), which complicated unambiguous read assignment and might essentially explain the low coverages across samples. *RHD* had, as expected, no coverage in the homozygous *RHD*01N.01* sample (S02). Its coverage also fell to around 10x in S04 and S05 due to the presence of a combination of *RHD* deletion (heterozygous *RHD*01N.01*) and *RHD* hybrid alleles (*RHD*03N.01* and *RHD*01EL.44,* respectively). Finally, S01 exhibited reduced *RHD* coverage, consistent with *RHD* hemizygosity. A similar trend was observed in S03, which is also hemizygous for *RHD*; however, its mean coverage (21.9x) was just above the outlier cutoff (19x) and therefore not classified as such. For male samples, genes located on the X chromosome were also expected to show less read coverage than those located on autosomes. Accordingly, S01, S02, S03 and S05, all samples from male individuals, exhibited reduced coverage for *GATA1* with outlier classification for S02 and S03. Reduced coverage below the 1$^{st}$ quartile was also observed for X-chromosomal *XK* gene in the male samples, but without hitting the outlier threshold. The two remaining X-chromosomal genes, *CD99* and adjacent *XG,* are fully and partially located in the pseudo-autosomal region (i.e. also on Y-chromosome) and, therefore, coverage is not expected to be reduced in males. However, *CD99* showed virtually absent coverage across all samples. Noteworthy, due to high homology between *CD99* located on X and Y-chromosome, only one *CD99* reference sequence (the one on the X-chromosome) was included in the FASTA file for adaptive sampling. However, not considering the Y-chromosome in the applied FASTA file unlikely played a role as there was no material *CD99* coverage in the female sample either. Upon further inspection of the BAM file using Integrative Genomics Viewer (IGV[24]; v2.17.4), we observed a sharp rise in coverage to expected levels starting from intron 3 of the *XG* gene about 11 kb downstream of *CD99*, which is exactly where the pseudo-autosomal region ends. To rule out bias from the T2T reference sequence, we aligned this region with the GRCh38 and GRCh37 reference sequences and found no significant differences that could account for this observation. In conclusion, *CD99* was excluded from downstream analyses.

#### 2.2 Variant Calling in C4A, C4B, CR1 and FCGR3B

Automatized variant calling in those 3 loci proved erroneous in our reference-based workflows I and II. A closer inspection of those loci showed homology over extended regions that surpassed the typical mean read length of ~10-23 kb, which resulted in ambiguous mapping against T2T.

In the case of *C4A* and *C4B*, the two genes are ~20 kb long and show >99% sequence identity across the entire gene[25], rendering gene-specific read mapping impossible without additional anchoring systems.

Unlike in GRCh38, the T2T reference genome shows an additional copy of a duplicated 81 kb segment in the *FCGR2/3* locus (CNR1=2), a relatively frequent CNV[26]. If samples present with a common *FCGR3B* gene dosage of two, i.e. one copy of CNR1 per haplotype, read coverage on T2T will drop to half, which is what we observed in several samples (Figure 2B). Proper variant calling would first require a CNV determination in our samples. Read assignment may otherwise not differentiate between real heterozygosity at a certain position or differences between the segments within the same haplotype.

*CR1* shows low copy repeats (LCRs), 18 kb repeat units, including 8 exons each, in some parts with more than 99% sequence similarity. Typically, SNV annotations in GRCh38 apply to only one unit, leaving large parts of the gene without annotations so far. Moreover, T2T shows fewer units than GRCh38 (39 exons instead of 47). To complicate things even further, adjacent gene *CR1L* includes also one of these homologous segments[27]. Unambiguous read assignment would require manual inspection or a tool targeted to such a gene system (Ref chinook).

#### 2.3 Allele assignments and phenotype predictions using RBCeq2

We detected consistent discrepancies for *FUT3* and *XK* alleles across all samples, which were due to a few missing null alleles in the software's data base and a nucleotide change error in ISBT allele tables for XK adopted in RBCeq2[28] (reference allele is c.1124C rather than c.1124G). Additional differences were observed for *GYPB* in S02 (RBCeq2 could not infer the hybrid allele, missing the MNS15 expression, while our manual prediction oversaw the altered MNS5 antigen due to the *GYPB*04.06* on both haplotypes) and in S03, where the splice site mutation c.270+5G>T was not picked up by the tool. An additional antigen encoded by the *SLC44A2* gene (CTL2.4) was not called by RBCeq2 either. At the time of analysis, RBCeq2 did not provide predictions for RH and CH/RG, which were manually added (supplemental Table 7).

#### 2.4 Rare variants verification results

Owing to the almost identical sequences of *C4A* and *C4B*, primer design specific to one or the other gene proved futile and those 32 variants (41 entries, supplemental Table 8) were omitted for replication analysis. We attempted Sanger sequencing for the two variants in *FCGR3B*, which both showed unequal allele frequencies in the variant calls of S04 and S02. Sanger sequencing also pointed to unusual allelic ratios, which could well be attributed to having amplified more than one segment, rather than to real heterozygosity, which is why we excluded these three entries. We could not confirm 8 variants in a repetitive part of *CR1* (exons 14 to 17), partly due to multiple failure of Sanger sequencing reactions. Variants assigned to exon 9 (N=3) could potentially be explained by mismappings as the T2T reference sequence of homologous exon 25 showed exactly these modifications. Similarly, the proposed variant in exon 19 was actually found to be represented in the reference sequence of homologous exon 9 of adjacent *CR1L*. Finally, a variant in exon 33 could be confirmed by Sanger sequencing. But since this exon is duplicated in GRCh38 (lifted over as rs113247278 in exon 41), we regarded all Sanger sequencing reactions of *CR1* as inconclusive and did not include the 13 *CR1* variants (16 entries) in the replication analysis.

#### 2.5 Sample-specific blood group alleles

In S01, we detected 130 exonic and flanking intronic variants (Table 1), among which one heterozygous frameshift mutation in *ABO* (the very common c.261delG; rs8176719 causing *ABO*O.01* alleles) and one heterozygous nonsense mutation in *FUT2* (c.461G>A; rs601338, resulting in the *FUT2*01N.02* allele). One common variant in the 5'UTR part of exon 3 in *ERMAP* was predicted to create a novel splice site 5 bp downstream of the existing one (Supp Table 5b), but this would not alter translation start site. Although splice site modifying prediction was minimal for rs72835417, this variant defines a null allele in *SID (SID*01N.02*). Apart from a weak allele in the PEL blood group system, we found the rare missense variant rs2534993131 in *AQP1* with a strong prediction of structural impact on Colton (CO) blood group antigens. Further variants with potentially deleterious effects mainly concerned *PIEZO1* for which the current allele collection is still rudimentary. Genotypes for 67 variants were available by MALDI-TOF MS and were all confirmed (supplemental Table 4)

For S02, from the 136 variants called within coding exons and flanking introns, we identified the same heterozygous frameshift mutation as in S01 (causing an *ABO*O.01* allele) and one heterozygous nonsense mutation in *GBGT1* (FORS blood group system) at coding position c.363G>T (rs35898523; p.Tyr121Ter), which, when combined with c.870G>A, results in *GBGT1*01N.04*. Although very rare, the phenotype is the same as in the reference allele *GBGT1*01N.01*, namely a non-functional enzyme. The exon 2 flanking variant rs12453819 in *B4GALNT2* intron 3 was predicted to form an alternative splice site, which would result in a three amino acid longer protein (in-frame). Novel missense variants with deleterious predictions were only found for genes where variant calling proved challenging or where hardly any variant have so far been reported in the ISBT allele tables (*C4A*, *C4B, CD44*, *ABCC1*,

*ABCC4*, *PIEZO1*). Most interesting was a low-frequent variant in *KLF1*, c.544T>C, where one affected allele may already cause the dominant In(Lu) phenotype. Genotype calls by MALDI-TOF MS differed for one variant out of 29. The *GYPA*01* vs. *GYPA*02* determining variant rs7682260 showed *GYPA*01* homozygosity in MALDI-TOF MS, but heterozygosity in Nanopore sequencing, a discrepancy explicable by the *GYP*401.02* hybrid allele.

Patient S03 showed a higher number of reliable short variant calls than the other samples, especially owing to an elevated number of synonymous SNVs (Table 1). The only frameshift variant was also detected in the *ABO* gene, but unlike in the other samples, we identified in S03 heterozygous c.1062delG, rs56392308, the *ABO*A2.01* defining indel. Additionally, we found the previously described nonsense mutation c.461G>A (rs601338) in the *FUT2* gene, this time in a homozygous state predicting a non-secretor phenotype. The second nonsense mutation, c.59T>G, was part of *GYPB*03N.04*, but its protein-terminating effect was abolished by adjacent c.60A>G affecting the same codon. Beside the aforementioned rs12124733 in *ERMAP* and rs12453819 in *B4GALNT2*, we found one further variant that was predicted to modify splicing: rare rs139511876 in *GYPB*, see main manuscript. From nonsynonymous SNVs with predicted pathogenic effects on protein function, most striking was rare c.695A>T (rs149648265) in *BCAM* on a *LU*02* background, a variant not yet reported in ISBT tables, but phenotypically most likely silent as in parallel of an inconspicuous *LU*02* allele. Of the 29 short variants genotyped with MALDI-TOF MS, we found no discrepancies with the results from Nanopore sequencing data.

In S04, we identified one frameshift and one nonsense mutation, both in heterozygous form. Again, it was a matter of the common indels rs8176719 and rs601338, characteristic for the *ABO*0.01* and *FUT2*01N.02* alleles. Two additional frameshift calls were faulty called due to the *RHD* hybrid allele. From the 12 reliable calls in flanking intronic regions, only rs12453819 in *B4GALNT2* predicted splice site consequences. We further found one weak allele in *ABCB6* and the known *KLF1*BGM12* that may not exhibit phenotypic consequences. A novel rare variant with high predicted impact on protein function was found in *GYPC* exon 4 (c.212T>C, rs115201071), a region so far devoid of allelic variability in the ISBT allele tables for the GE blood group. The 29 genotypes called by MALDI-TOF MS could all be confirmed.

Finally for S05, we detected *ABO*O.01* causing frameshift variant c.261delG in homozygous form. In one allele, it was combined with nonsense (but in this case untranslated) c.542G>A (rs55727303). We also found the nonsense mutation c.461G>A on one *FUT2* allele. The detected *RHD* variants proved to a large part hybrid-related, including one frameshift mutation. An exception was synonymous c.504C>T lying on *RHCE*-originating exon 4, which we could confirm by Sanger sequencing. As already in 3 other samples, rs12453819 in *B4GALNT2* was also detected in S05. We identified two novel nonsynonymous *BCAM* variants, low-frequent c.586G>A (MAF=~0.02) on a *LU*01* and rare c.1766C>T (MAF=~0.003) on a *LU*02* background, the latter showing a higher pathogenic risk score. We also detected one *JK*02N.17* allele, but impact of synonymous c.810G>A to cause a null allele is questionable[29]. We did not find any discrepancies between the 29 called variants by MALDI-TOF MS and our in-house variant calling workflow based on Nanopore sequencing.

### Section 3: Discussion

#### 3.1 Improvement of reference-based variant calling

Usage of single reference genomes for variant calling, and especially SV calling, has faced increasing criticism due to its inability to capture the full spectrum of human genetic diversity. For instance, while comparing high coverage sequenced dataset of 910 individuals from African descents, Sherman et al.[30] found 296.5 Mb of novel DNA sequences missing from the GRCh38 reference genome. Hence, the African pangenome reference they created contained ~10% more DNA than the at that time current "linear" human reference genome. Although using the latest T2T reference genome improved genomic analyses[31] by integrating the remaining 8% of "genomic dark matter", correcting thousands of structural errors and unlocking the most complex regions[32], no single genome can represent the global genetic diversity. To address this limitation, the adoption of pangenomic references has gained significant momentum in recent years because it offers considerable advantages over traditional single-reference approaches. For instance, it has been shown to significantly reduce both false-positive and false-negative variant calls, particularly at complex genomic loci[33,34].

Behera et al.[35] proposed a methodology known as Dynamic Read Analysis for Genomics (DRAGEN) to facilitate the identification of various genomic variant types for Illumina short read WGS data. The DRAGEN variant calling pipeline enhances the precision of variant detection through multigenome mapping utilizing pangenome references and machine learning-based filtering for variant calling. Additionally, given the challenges associated with genotyping certain critical genes—attributable to paralogous genes or repetitive regions—DRAGEN integrates nine specialized variant callers to ensure accurate genotyping of clinically significant genes, one of which is specifically designed for *RH*[35].

Despite the substantial advancements pangenomic references have made (20), several challenges remain. Their use require specialized tools, lack automated pipelines, and demand high computational resources. They also complicate data alignment, genomic coordinate representation, and software compatibility, necessitating new bioinformatics tools. Managing ISBT tables and reference annotations would also require advanced data frameworks when coupled with pangenome references.

### Section 4: Supplemental Figures

**Figure S1: Circos plots showing read coverage across the genome for samples S02 (A), S03 (B), S04 (C) and S05 (D)**. Genes of interest are indicated according to chromosomal positions around the outer circle (n=64). Mean coverages across genome (100 kb windows) is shown by black lines; mean coverages in ROI are further highlighted in red. Circles of dashed lines represent mean read coverage within (black) and outside (white) ROI. The corresponding circos plots for S01 is shown in Figure 2A.

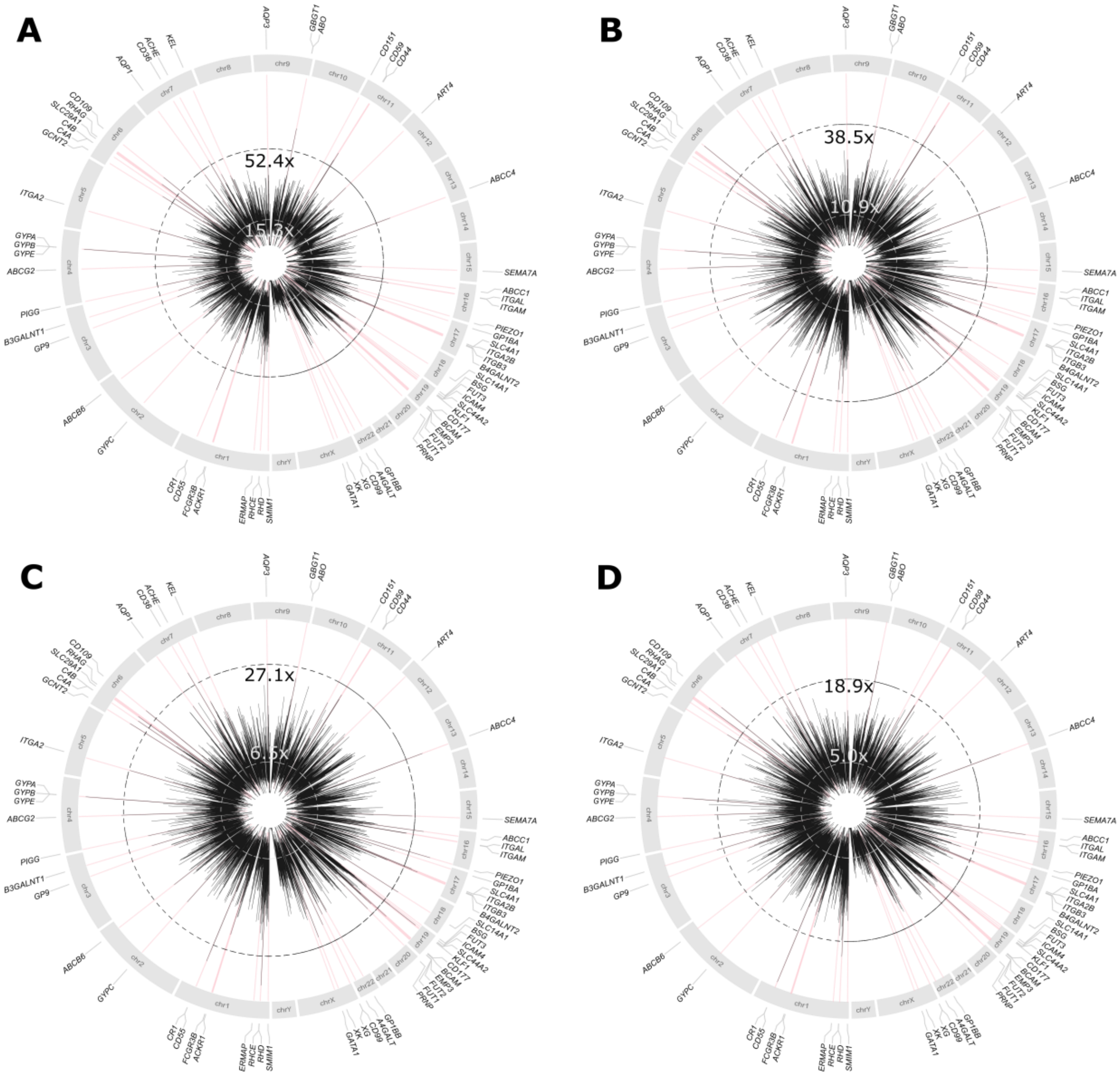
